# Policy Gaps in Type 1 Diabetes: A UK Audit of National Governing Bodies

**DOI:** 10.1101/2025.08.05.25333018

**Authors:** Mohammed Abdel-Magid, Emma J. Cockcroft, Chris Bright, Robert H. Mann, Richard Pulsford, Robert C. Andrews

**Affiliations:** Department of Endocrinology and Diabetes, North Bristol NHS Trust, Bristol, UK; University of Exeter Medical School, Exeter, UK; Breakthrough T1D, UK; Department of Public Health and Sport Sciences, University of Exeter, Exeter, UK; Podium Analytics, London, UK; Exeter NIHR Clinical Research Facility, Exeter, UK; NIHR Exeter Biomedical Research Centre (BRC), Exeter, UK

**Keywords:** Diabetes, chronic diseases, health policy, physical activity, inclusion, participation

## Abstract

**Aims:** National Governing Bodies (NGBs) for sport in the United Kingdom (UK) are responsible for setting standards and ensuring safe, inclusive participation for all, including people with Type 1 Diabetes (T1D). This audit systematically assessed the extent and quality of policy provision among UK-based NGBs, focusing on support for athletes with T1D. The findings were compared with epilepsy, asthma, and allergies to evaluate equity and consistency in safeguarding and inclusion practices.

**Methods:** *Design:* A systematic policy audit of UK sport NGBs to assess the presence and content of policies addressing athletes with T1D, asthma, epilepsy, and allergies.

*Data Sources:* Policy documents were accessed through public materials on NGB websites. NGBs were contacted by email to confirm the existence of relevant policies, and to request any documents not currently included on their website.

*Eligibility Criteria:* Sports were included if their NGB had a publicly accessible website.

*Results:* Of 185 NGBs, 20 (11%) had policy documents mentioning diabetes, 14 (7.7%) asthma, 12 (6.6%) epilepsy, and 4 (2.2%) allergies. Of the 20 NGBs with documents mentioning diabetes, only 4 had dedicated diabetes policies. The remaining 16 referred to diabetes in broader documents.

*Conclusion:* Despite increasing emphasis on inclusion in sport, few NGBs have clear policies to support individuals with diabetes. This is also the case for asthma, epilepsy, and allergies. We recommend the development of national policies for diabetes, which can be adapted by individual sporting bodies.

**Equity, diversity, and inclusion statement:** The author group is gender balanced and consists of junior and senior researchers from a range of disciplines and ethnic backgrounds, based in the UK and one African country. Our study population included sports organisations in the UK, covering a diverse range of sociodemographic backgrounds and populations; although findings may not be generalisable to settings with fewer resources.

**What is already known?:**

- Regular physical activity helps to improve control, boost wellbeing, and reduces long-term complications in people with diabetes, however many people with diabetes are not meeting recommended levels of physical activity.
- National Governing Bodies play a pivotal role in enabling and safeguarding participation of those with diabetes in sport.
- Coaches frequently lack the essential training and expertise required to support athletes living with diabetes.

**What this study has found?:**

- Out of 185 National Governing Bodies in the United Kingdom, only four (2%) have dedicated policies to support people with diabetes.
- Provision for athletes with epilepsy, asthma, and allergies is similarly scarce.

**What are the implications of the study?:**

- We recommend a national diabetes sporting policy is developed that can be modified by National Governing Bodies to reflect the needs of their specific sport.
- Similar policies should also be developed for other common chronic health conditions, including epilepsy, asthma, and allergies.

## INTRODUCTION

Type 1 diabetes (T1D) is a Chronic Health Condition (CHC) with significant healthcare needs. It is associated with an increased risk of coronary artery disease, stroke, renal failure, retinopathy, and premature death^1,2^. Recent estimates indicate that 9.5 million individuals worldwide are living with T1D, a 13% increase from 8.4 million in 2021. From this total, approximately 1.0 million are aged 0–14 years and 0.8 million are aged 15–19 years ^1^. Within the United Kingdom (UK), approximately 341,000 individuals were living with T1D as of 2024–2025, including about 30,000 children and young people aged 0–18 years, representing one of the highest prevalence figures in Europe ^1,3^. Effective management of T1D necessitates rigorous self-management, multiple daily insulin injections, frequent glucose monitoring, and disciplined dietary and activity regulation.

Despite technological advancements in diabetes care, optimal glycaemic control remains difficult to achieve for many Physical activity (PA) is important in T1D management, along with insulin and nutrition. Regular PA improves glycaemic control, reduces insulin requirements, and supports cardiovascular health, minimising long-term complications ^2^. Beyond physiological benefits, regular PA during childhood and adolescence facilitates healthy physical and psychological development, fosters self-confidence, enhances mood, and delivers social advantages ^4^.

Recognising these benefits, international and UK PA guidelines recommend adults achieve at least 150 minutes of moderate-to-vigorous intensity PA each week, while children and adolescents engage in a minimum of 60 minutes of moderate-to-vigorous activity daily ^5^.

Despite this, people with T1D consistently report lower levels of PA compared to their peers ^6^. This can be attributed to internal and external factors, including lack of motivation; fear of hypoglycaemia; negative emotions (e.g., fear, low mood); uncertainty about safe PA practices/locations; and/or insufficient support from family and peers ^7^. Participation in PA, therefore, requires help from sports organisations, coaches, and healthcare professionals to ensure equitable access and maximise health and social advantages for people with CHCs.

In the UK, National Governing Bodies (NGBs) regulate sports at a national level and occupy a central role in determining inclusion, as their policies delineate which individuals are permitted to participate and under what conditions. NGBs are central in developing, implementing, and monitoring policies that support the health and wellbeing of participants ^8^, ensuring policies are inclusive to people with CHCs and have provisions to ensure the safety of individuals during participation – as a fundamental duty of care^9^. However, there have been news reports on participants from different sports (e.g., football, cricket) who died from T1D-related complications, most notably Diabetes Ketoacidosis (DKA) and hypoglycaemia ^10,11^. Many coaches lack the knowledge and skills to support individuals with T1D, and can be complicit in generating or enhancing stigma experiences by those with T1D^12^. NGBs could implement policies that provide tailored advice and safety protocols for people with T1D, as recommended by the World Health Organization (WHO) ^13^. This approach aligns with the UK Equality Act 2010, as T1D can be recognised as a disability; and the United Nations Convention on the Rights of Disabled People (CRPD) Article 30, which states all disabled people have the right to equitable sport, leisure and recreation. Both legislations justify the need for additional support and protection from discrimination.

To date, no research has evaluated the prevalence or implementation of T1D policies among UK-based NGBs. This audit aims to assess the extent to which NGBs have established and made publicly available policies that support individuals with T1D, and to compare this to NGB policies for asthma, epilepsy and allergies. It examines the content of these policies to determine whether they address the needs of sport participants, both recreational and competitive.

## METHODS

This audit was guided by the first 5 steps of the methodological framework outlined by Arksey and O’Malley and informed by Levac and colleagues ^14,15^. This approach provides a rigorous, transparent approach that explicitly accommodates grey literature and policy documents, which were central to this review.” The stages were as follows:

***Stage 1:*** *Defining the Research Question* – We specified the overarching questions as: (1) how many UK NGBs have publicly available policies to support individuals with T1D and other CHCs?; and (2) what is the content of these policies with respect to participation and safety?
***Stage 2:*** *Identifying Policies – NGB websites were manually searched (July–August 2024) by three reviewers (MA-M, EC, RA) using the terms ‘diabetes’, ‘asthma’, ‘epilepsy, and ‘allergy’. Each NGB was then contacted by email to confirm the existence of relevant policies and to request documents not available online; a reminder email was sent after two weeks to non-responders*.
***Stage 3:*** *Inclusion/Exclusion criteria – All NGBs recognised by the UK Sports Councils and listed by Sport England* ^16^ *were eligible (n = 185). NGBs were included if they had a publicly accessible website. Documents were included if they: (a) originated from an eligible NGB; and (b) contained any reference to T1D, asthma, epilepsy, and/or allergies*.
***Stage 4:*** *Charting the Data – For each NGB we extracted: (1) whether official policies existed for diabetes, asthma, epilepsy, and/or allergies; and (2) the exact wording of relevant policy statements where a dedicated policy was not available. Offline copies of health declaration forms were downloaded. If separate policies were not found online, broader documents were reviewed for references to these conditions. This included safeguarding policies, risk assessment policies, and event guidelines. Where relevant references were found, the information was recorded. All data were systematically documented in Microsoft Excel 2019 to allow for comparison and analysis across NGBs*.
***Stage 5:*** *Collating, Summarising, and Reporting – We generated descriptive counts and proportions of NGBs with relevant policies and summarised the policy content narratively, focusing on the extent to which documents addressed inclusion, safety, and practical guidance for athletes with CHCs*.

In line with Arksey and O’Malley’s original framework ^14,15^, the consultation stage (Stage 5) is considered optional. For this audit, a consultation stage was not undertaken because the primary objective was to systematically map existing, publicly available NGB policy documents rather than to develop or refine policy content in partnership with stakeholders. Informal input from clinical and sport-policy experts within the authorship group informed interpretation of the findings, but no separate structured consultation phase was conducted.

## RESULTS

Of the 185 UK-based NGBs identified, 184 had working websites. Of these, 176 were successfully contacted, 137 via email and 39 through online contact forms. The eight remaining NBGs were not contactable due to lack of email or online contact form (n = 2) or bounced email/failed contact form (n = 6). Thirty-three of the successfully contacted NGBs (19%) responded to our enquiries (Figure 1).

**Figure 1:**
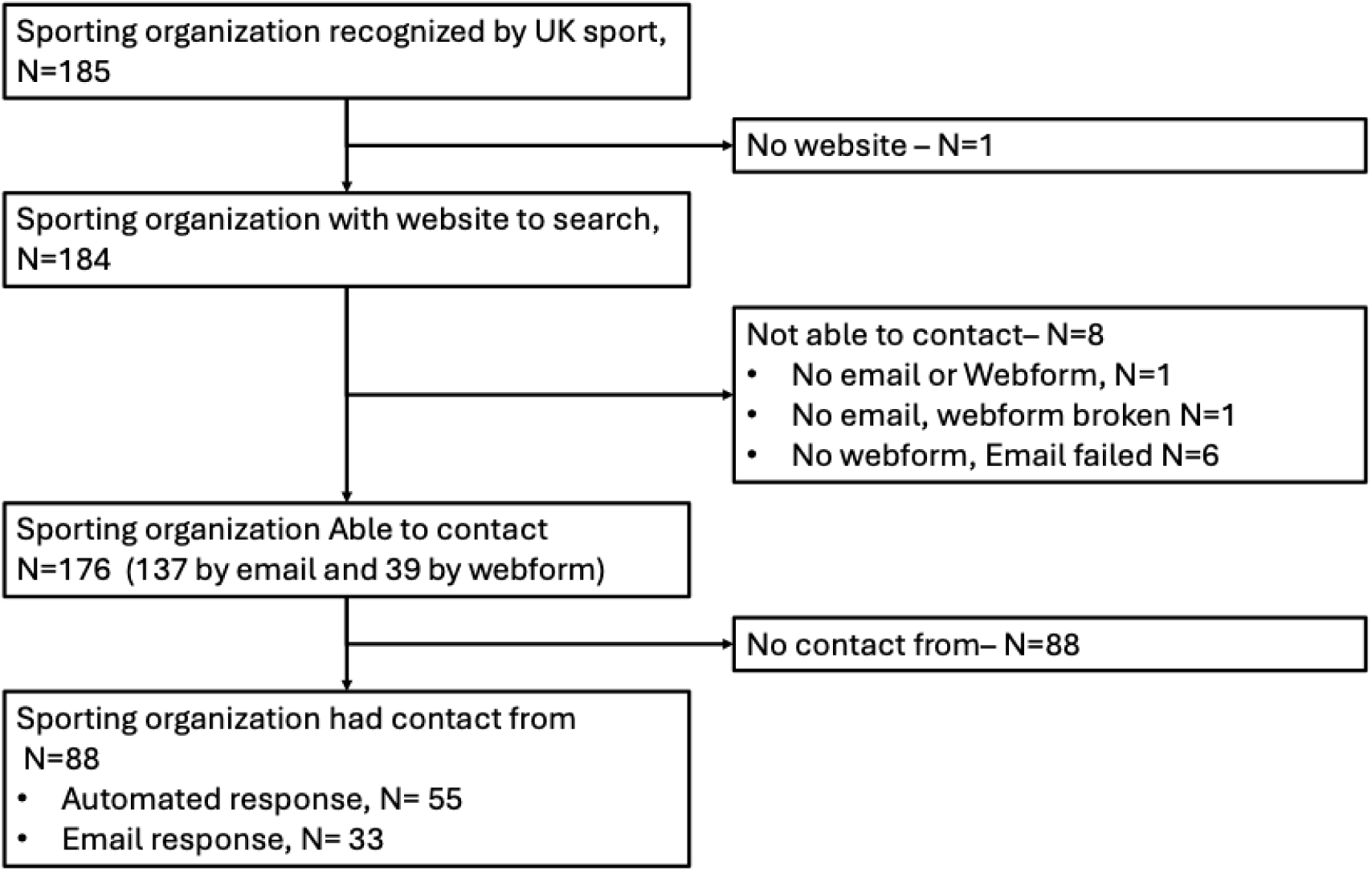
Flow diagram of study information. The websites of 185 UK sports organisations were screened for relevant policy documents. One organisation did not have a website. We were able to contact 176 organisations to confirm the presence of policies and to request any unpublished documents; eight could not be contacted due to issues with their webforms or email addresses. A reminder email was sent after two weeks to non-responders. In total, we received 88 responses — 55 were automated replies and 33 were written email responses

### Overview of published policies

An overview of the number of NGBs with policies relating to different is shown in Figure 2. Table 1 summarises the content of diabetes and CHC policies.

**Figure 2:**
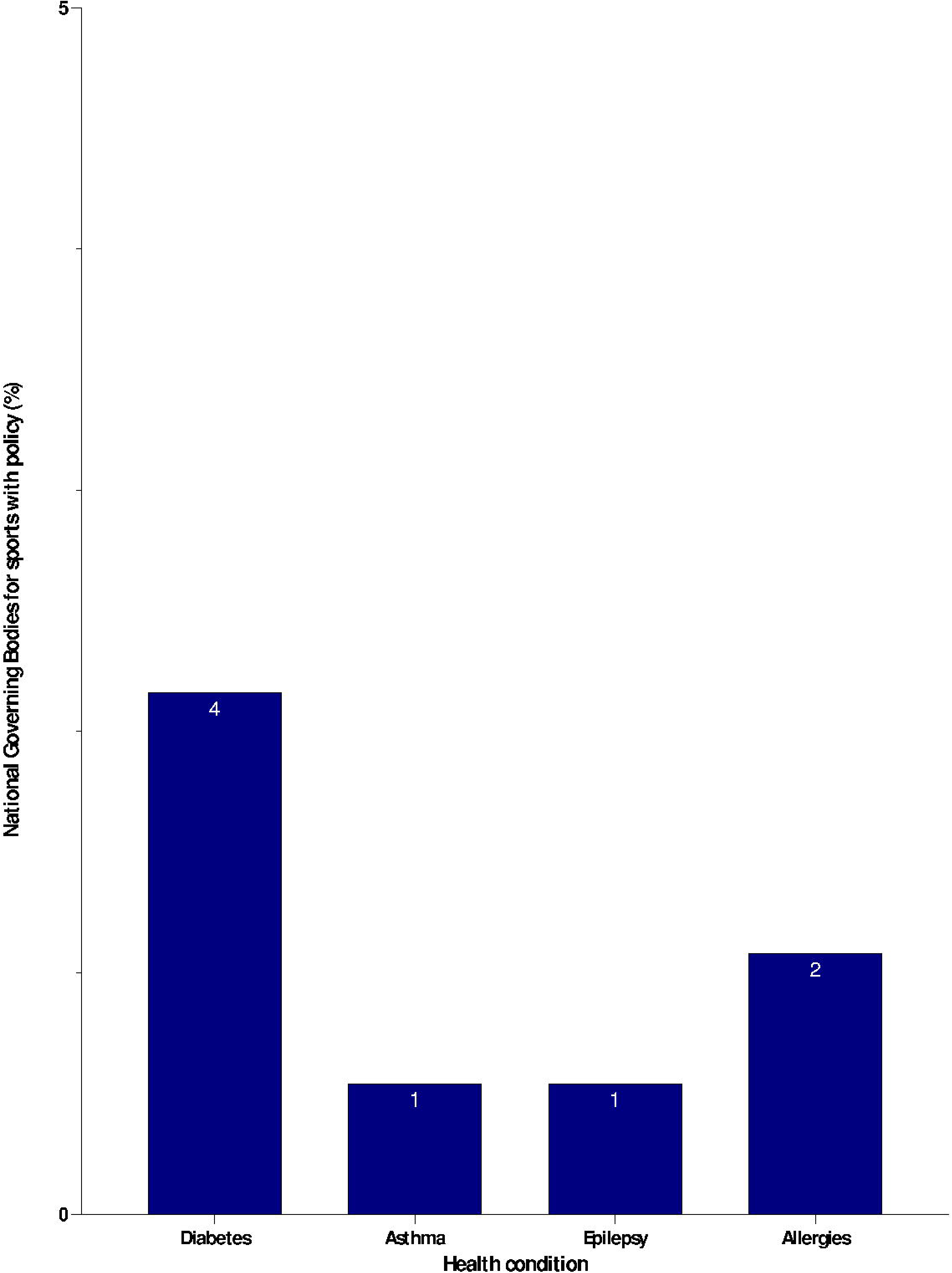
Number of National sporting bodies with a policy on Type 1 diabetes, asthma, and epilepsy. Prevalence of health condition-specific policies among national governing bodies (n=185). Data represents the percentage of NGBs with policies or guidance specific to each health condition. Diabetes (2.2%), allergies (1.1%), asthma (0.5%), and epilepsy (0.5%).

**Table 1.**
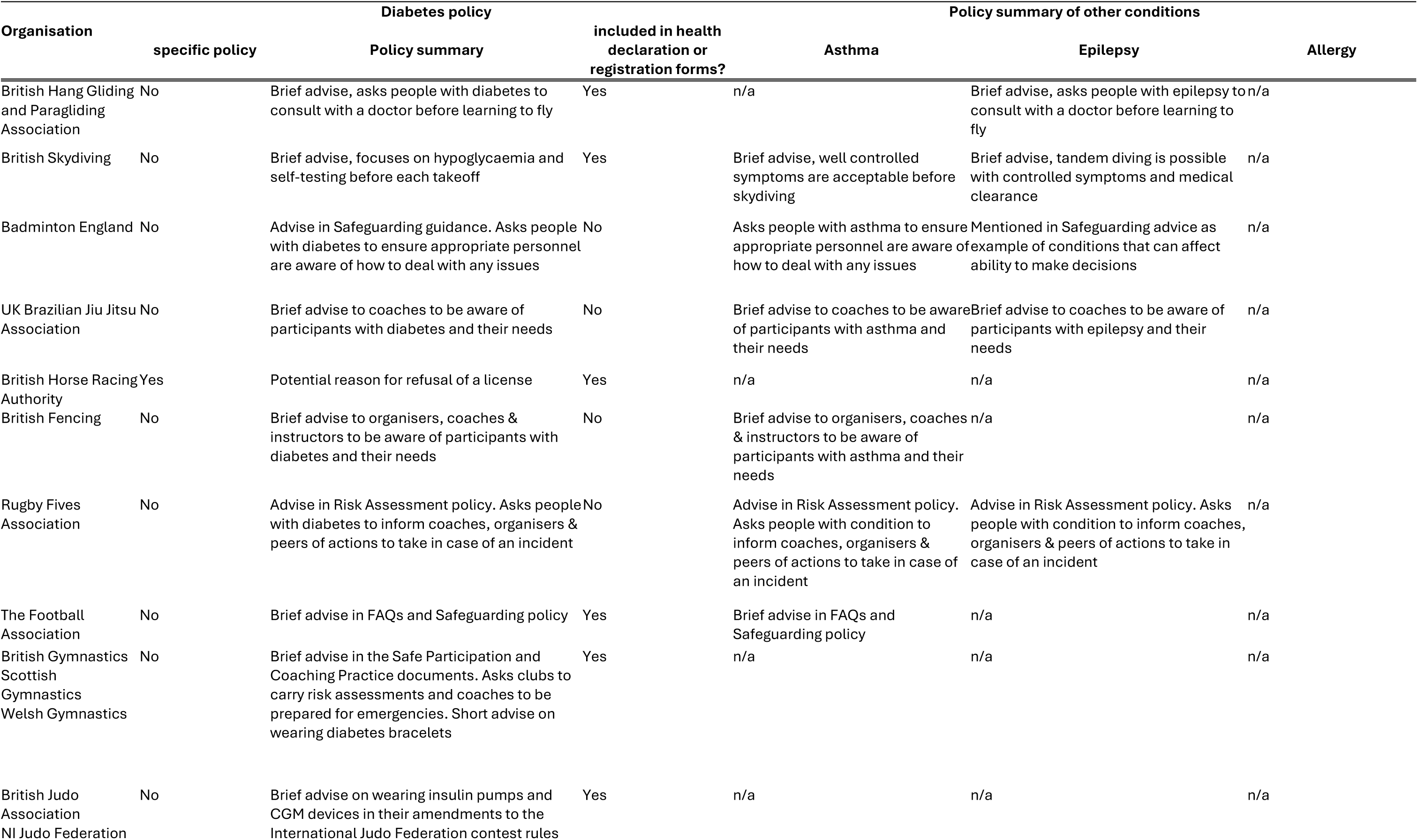
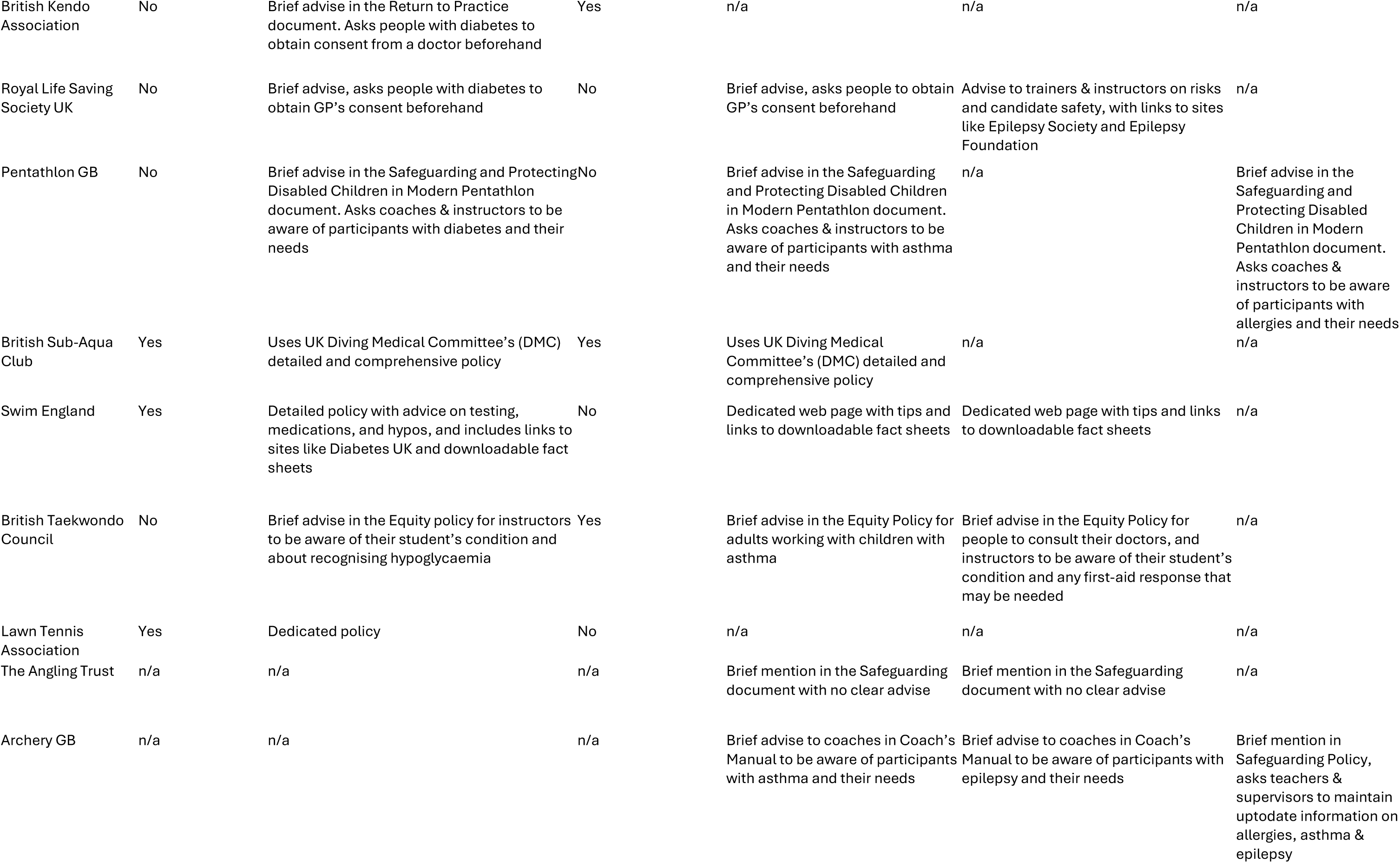
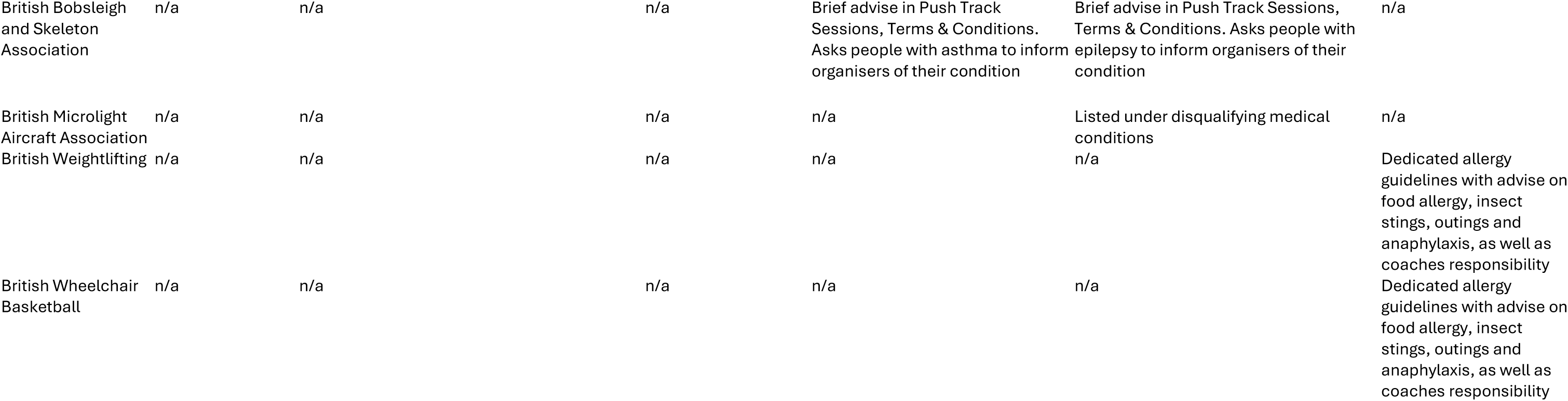
NGB’s with policies relating to diabetes, asthma, epilepsy, and allergies.

### Diabetes

Out of 185 NGBs, only 20 (11%) mentioned diabetes (Type 1 or Type 2) on their websites or in policy documents. Only 4 (2%) had dedicated policy documents to support people with diabetes: (1) British Horse Racing Authority (BHRA); (2) British Sub-Aqua Club (BSAC); (3) Swim England (SE); and (4) Lawn Tennis Association (LTA).

The remaining 16 (9%) NGBs included short diabetes advice in other documents, such as: (1) equality, diversity and inclusion policies (n = 1); (2) safeguarding policies (n = 5); (3) event guidance documents (n = 9); and (4) risk assessment and safety policies (n = 5). Eleven NGBs asked about diabetes in their health registration/declaration forms. None of the 20 NGBs specified whether insulin must be declared under a Therapeutic Use Exemption (TUE). However, two organisations - British Judo and Northern Ireland Judo - included brief guidance on the use of insulin pumps within their event documentation. Details of the documents in which diabetes was mentioned are summarised in Figure 3.

**Figure 3:**
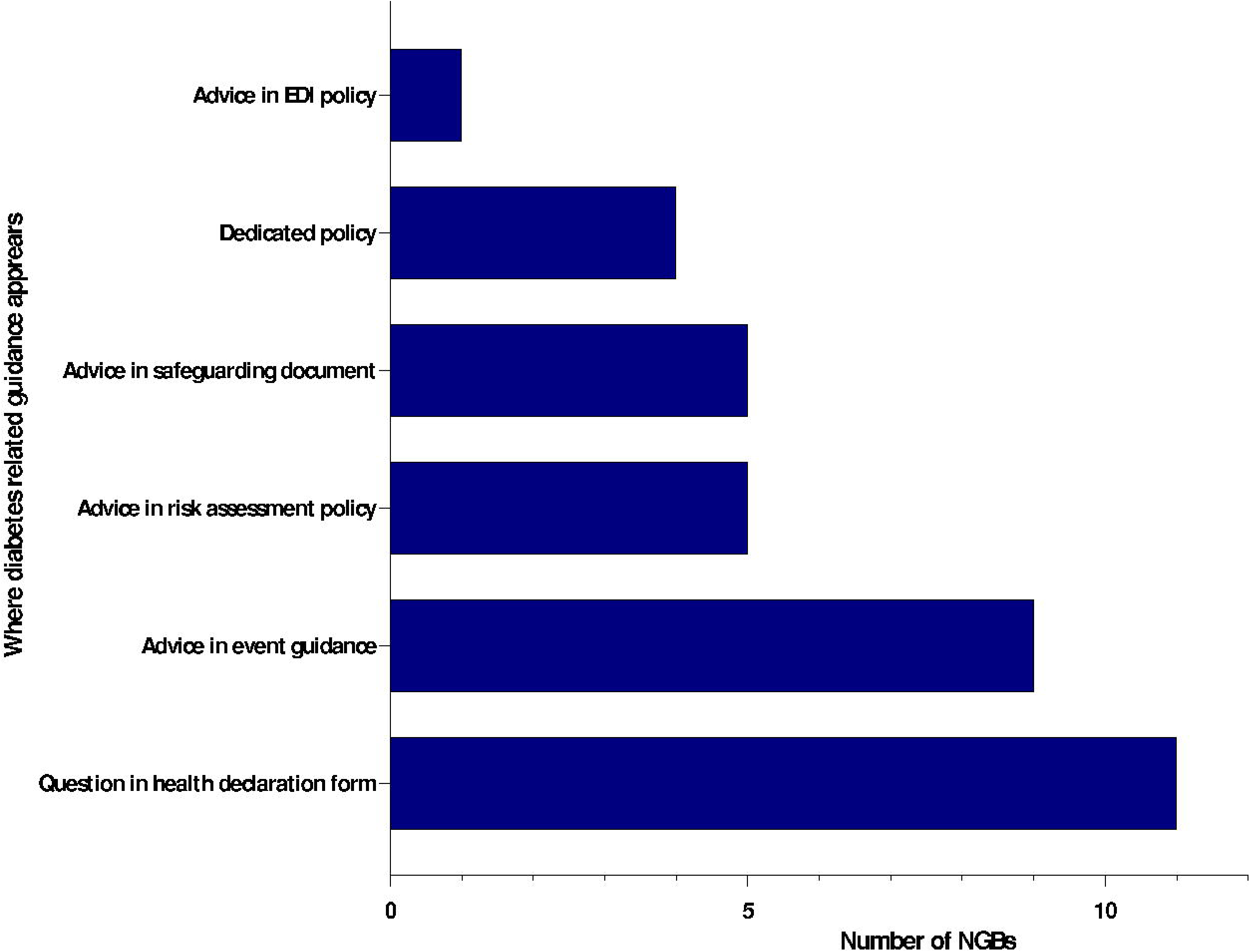
Types of documents in which diabetes was mentioned. Types of documents in which diabetes was mentioned. Only four organisations had dedicated diabetes policies. Three of these, along with an additional sixteen organisations, included diabetes-related guidance within other documents. EDI: Equality, Diversity & Inclusion

### Asthma

Fourteen NGBs (8%) mentioned asthma, but only one (BSAC) had a dedicated policy and one (SE) had a guidance webpage. The other 12 included short advice in other policy documents.

### Epilepsy

Twelve NGBs (7%) mentioned epilepsy, but only one (SE) had a dedicated guidance webpage. All other mentions were in different policy documents.

### Allergies

Four NGBs (2%) mentioned allergies, and only two (British Weightlifting and British Wheelchair Basketball) had dedicated policies. The others were brief mentions in other policy documents.

### Review of NGB diabetes-related advice and policy wording

Most diabetes policies focused on equipping athletes with the knowledge and skills to manage their diabetes, and emphasised the involvement of appropriate personnel (e.g., coaches, event organisers). For example, the British Hang Gliding and Paragliding Association’s advice on their “Learn to Fly” page reads:

> *“If you suffer from any medical condition such as epilepsy, … or diabetes you should ask your Doctor’s advice.”*

Similarly, Badminton England advises in their Safeguarding Guidance document:

> *“If any of the group have asthma, diabetes, severe allergies or any other condition, ensure that appropriate personnel travelling with the group are aware of how to deal with any given situation.”*

Several documents recommended getting medical clearance before participating in PA. For example, the British Kendo Association’s Return to Practice document states:

> *“Who Should Not Participate/Attend?*

> *People with underlying conditions such as diabetes, heart disease, …, etc. If people with these conditions intend to participate consent from a doctor should be obtained beforehand.”*

Other policies advised coaches and instructors to remain vigilant for participants with diabetes, recognise signs of hypoglycaemia, and be prepared for emergencies, usually without specific detail. Examples include UK Brazilian Jiu Jitsu Association’s Code of Conduct, which states:

> *“The coach should also be aware if participants have long-term medical conditions such as epilepsy, diabetes or asthma. As a coach you should consider the participants’ specific needs; what they can do as well as what they cannot do and the barriers they face to participation. There are certain medical conditions that may inhibit participation; coaches are advised to seek professional advice … before attempting to devise a training programme.”*

British Taekwondo Council’s Equity policy stated:

> *“Instructors working with students with diabetes should be aware: (1) of the symptoms associated with the onset of hypoglycaemia (2) that a person with diabetes may carry with them a bag containing a blood glucose testing kit, food, glucose tablets, drinks etc, which he/she should be allowed to use as and when necessary”*

Among the four NGBs with dedicated diabetes policies, the covered topics included: (1) fitness to exercise (BHRA and BSAC); (2) risks of hypoglycaemia and hyperglycaemia (BSAC, SE, LTA); (3) checking blood glucose throughout exercise (BSAC, BHRA, SE); (4) emergency treatment of hypoglycaemia (BSAC, SE, LTA); (5) nutrition and hydration (BSAC, LTA); (6) safety measures during PA (BSAC, BHRA, SE, LTA); and (7) adjustments to diabetes treatment (SE).

These examples highlight a general focus on risk awareness and medical oversight but lack detailed and practical guidance for coaches and event organisers.

## DISCUSSION

This audit highlights a significant policy deficit within UK sport for individuals with T1D. Of 185 NGBs, only 20 (11%) referenced diabetes in their websites or policy documents; of these, 4 NGBs (2%) possessed dedicated policy documents to support people with diabetes. Consequently, 165 (89%) NGBs provide no guidance whatsoever regarding T1D. This marked absence of dedicated policies persists despite explicit WHO recommendations for safeguarding individuals with T1D in PA settings ^13^.

The lack of tailored guidance is concerning given the central role NGBs occupy in establishing national standards, directing practice, and ensuring the welfare of participants with T1D. By not prioritising this demographic, most NGBs forfeit opportunities to eliminate participation barriers, improve health outcomes, and uphold their legal and ethical obligations ^17^. In a recent Breakthrough T1D (formerly Juvenile Diabetes Research Foundation) survey, one third of respondents identified lack of education among coaches and organisers as a barrier to PA, while nearly one quarter reported stigma and negative remarks ^18^. This policy gap may partly explain these findings.

National policy must be robust to transform PA culture at all levels. When NGBs set inclusion standards, local clubs and coaches follow. Conversely, policy absence results in limited awareness and missed opportunities to support those with additional health needs. Sport England’s local delivery pilots show that national strategy can influence local outcomes and reduce inactivity ^19^. Psychological safety empowers staff, coaches, and participants to raise concerns and criticise behaviours that may compromise wellbeing, creating a culture of inclusiveness. As Knott and colleagues highlight, safe and ethical sport require more than policies; they demand a shared, relational culture where dialogue and reflection are normalised and whistleblowing is protected ^20^.

Limited document accessibility is another concern. T1D mentions were often hidden in safeguarding or EDI policies and required specific keyword searches. Effective policies must be labelled and accessible from NGB homepages. Without visibility, implementation becomes unlikely.

Clear policy has proven benefits across sectors. As Skiba notes, clear business policies ensure legal compliance, guide staff, set behaviour and performance expectations, and improve productivity ^21^. These principles apply to NGBs, where accessible policies give staff, volunteers, and participants clear guidance, building confidence, consistency, and compliance. This is important in sport, where staff and volunteers may have varying levels of experience and training. Policies that are difficult to find, complex, or ambiguous may be misunderstood or ignored, undermining compliance and the inclusive culture they promote. Ultimately, accessible policies are not just a compliance requirement; they are essential for governance, inclusion, and organisational success.

The finding that asthma, epilepsy, and allergies have similar policy gaps suggests a systemic issue affecting CHCs, not only T1D. To illustrate how policy attention can differ across marginalised groups, we compared this gap with transgender policies, acknowledging that both people with CHCs and those whose gender identity differs from that recorded at birth face major barriers to participation.^22,23^. It is encouraging that 54 of 185 (29%) NGBs have transgender inclusion policies, aligning with international frameworks such as the International Olympic Committee’s position on fairness and non-discrimination ^24^. This disparity may be explained by the stronger legal, structural, and political drivers for transgender policy development. The Equality Act 2010 explicitly protects “gender reassignment,” placing clear statutory obligations on sports organisations, whereas CHCs fall under broader disability provisions with less compliance pressure. Moreover, the Sports Councils’ Equality Group issued detailed national guidance on transgender inclusion in 2021, providing NGBs with ready-made frameworks and legal clarity—resources not available for CHCs, where guidance remains fragmented across health organisations. Finally, transgender participation has attracted sustained political and media attention, prompting high-level government engagement, whereas CHCs in sport have only recently begun to receive comparable visibility. Given there are more people in the UK with CHCs than those who are transgender, and they are proportionately being excluded from inclusive policy, there is a clear policy gap that requires immediate action.

To close this policy gap, Breakthrough T1D and others have underscored the value of co-producing resources with individuals with lived experience of CHCs. Their collaborations with NGBs, including the Football Association of Wales and the LTA, show how integrating community insight can lead to practical outcomes like condition-specific guidelines for coaches and athletes that improve support and reduce stigma ^25^. Expert recommendations and international frameworks recommend clear, practical, and inclusive policies for athletes and support staff. Nationally endorsed sport-specific CHC policy templates could help ensure consistency, facilitate uptake across disciplines, and uphold NGBs’ equality obligations.

According to the National Athletic Trainers’ Association (NATA) and the International Diabetes Federation (IDF), every athlete with diabetes should have a care plan that covers glucose monitoring, recognition and treatment of hypo- and hyperglycaemia, nutritional advice, insulin dose modification, travel, footcare, and compliance with anti-doping regulations, such as the World Anti-Doping Agency (WADA) ^26^. A dedicated diabetes sports policy should include all the above and apply to both recreational and professional athletes.^27^ However, our findings show that most T1D advice is embedded within risk assessments, safety, or equality, diversity and inclusion (EDI) policies, and often requires individuals to obtain medical clearance and inform coaches of their condition. The term ‘incident’ was frequently undefined, and even the four dedicated diabetes policies lacked guidance on insulin or insulin pump management. For example, in the Judo association’s event guidance, insulin pumps were banned in competition without offering alternatives or directing athletes to further advice. This is not inclusion; it is exclusion by omission.

It is also essential to avoid unnecessary barriers. Medical clearance is not needed for people with T1D doing moderate exercise, unless they have heart disease, plan high-intensity or endurance activities, or take part in sports requiring clearance, like scuba diving ^28^. However, semi-professional and professional athletes benefit from consulting a diabetes specialist, particularly one with expertise in sport or exercise physiology. Managing T1D in this context involves personalised adjustments to insulin regimens, carbohydrate intake, and glucose monitoring to minimise the risk of hypoglycaemia and hyperglycaemia ^29^.

Some centres offer “diabetes sports clinics” to educate athletes on new technologies, travel, competition, and injury recovery. These clinics also help submit TUE forms. Diabetes-specific sports policies should incorporate this expert advice and extend it to coaches, event organisers, and other relevant personnel to improve safety, inform decision-making, and promote diabetes inclusion in sport at all levels.

### Research/Policy Implications

This audit demonstrates that most UK-based NGBs lack clear, accessible policies to support athletes with diabetes and other CHCs. This has implications for policy and research. At policy level, NGBs must create dedicated, visible, and practical guidance for these groups, rather than relying on brief references hidden within safeguarding or diversity documents. Making policies prominent and comprehensive helps coaches and clubs to protect the health and wellbeing of participants with CHCs, and achieve their legal and ethical obligations under the Equality Act 2010.

Looking ahead, this issue is only set to grow in importance. The Health Foundation projects that by 2040, nearly one in five adults in England will be living with major illness, with diabetes rising the most ^30^. Failure to act now to embed inclusive sport policies will worsen future inequalities in access to PA, especially as more people are living with complex health needs.

Future research should explore whether better policy implementation increases participation, improves safety, and reduces stigma for these athletes. Understanding barriers to policy implementation and dissemination within sports organisations is equally important. Further qualitative research may help clarify the views and needs of athletes, coaches, and administrators. All these areas must improve for people with CHCs to benefit from safe, inclusive sports.

### Limitations

This audit offers a novel and comprehensive assessment of publicly available health-related policies among UK-based NGBs regarding diabetes, asthma, epilepsy, and allergies. Nevertheless, several limitations should be noted.

First, only UK-based NGBs were included. The findings may not apply to sports organisations in other countries where policy frameworks, resources, or digital access differ. International comparative studies are needed to address this gap.

Second, the analysis included only online and publicly accessible documents. Some NGBs may have relevant internal or unpublished materials that were not identified. Not all NGBs responded to our enquiries; therefore, lack of visible policy does not necessarily imply no policy.

Lastly, this review did not assess policy implementation or impact. Policy existence does not guarantee its use or efficacy. Further research should examine how policies are enacted at grassroots and elite levels, and whether they improve inclusion, safety, and participant outcomes.

## Conclusion

This audit highlights a significant gap in UK sport policy for people with T1D and other CHCs. There is an urgent need for clear, evidence-based guidance that NGBs can adopt to ensure safe and inclusive participation. This should be partnered with strengths of the Equality Act 2010 for CHC and greater political pressure. Without such actions unnecessary barriers will persist, making sport less accessible and equitable than it should be.

## Data Availability

The datasets generated and/or analysed during the current study are not publicly available but are available from the corresponding author on reasonable request.

## Acknowledgements

This work forms part of the research portfolio of Podium Analytics, a charity with the mission of More Sport Less Injury (podiumanalytics.org). This study was supported by the National Institute for Health and Care Research Exeter Biomedical Research Centre and National Institute for Health and Care Research Exeter Clinical Research Facility. The views expressed are those of the authors and not necessarily those of the NIHR or the Department of Health and Social Care.

This research received no external funding.

